# Microarray-based detection of antibodies against SARS-CoV-2 proteins, common respiratory viruses and type I interferons

**DOI:** 10.1101/2021.12.13.21267509

**Authors:** Elena Savvateeva, Marina Filippova, Vladimir Valuev-Elliston, Nurana Nuralieva, Marina Yukina, Ekaterina Troshina, Vladimir Baklaushev, Alexander Ivanov, Dmitry Gryadunov

## Abstract

A microarray-based assay to detect IgG and IgM antibodies against betacoronaviruses (SARS-CoV-2, SARS, MERS, OC43, and HKU1), other respiratory viruses and type I interferons (IFN-Is) was developed. This multiplex assay was applied to track antibody cross-reactivity due to previous contact with similar viruses and to identify antibodies against IFN-Is as the markers for severe COVID-19. In total, 278 serum samples from convalescent plasma donors, COVID-19 patients in the intensive care unit (ICU) and patients who recovered from mild/moderate COVID-19, vaccine recipients, prepandemic and pandemic patients with autoimmune endocrine disorders, and a heterogeneous prepandemic cohort including healthy individuals and chronically ill patients were analyzed. The anti-SARS-CoV-2 microarray results agreed well with the ELISA results. Regarding ICU patients, autoantibodies against IFN-Is were detected in 10.5% of samples, and 10.5% of samples were found to simultaneously contain IgM antibodies against more than two different viruses. Cross-reactivity between IgG against the SARS-CoV-2 nucleocapsid and IgG against the OC43 and HKU1 spike proteins was observed, resulting in positive signals for the SARS-CoV-2 nucleocapsid in prepandemic samples from patients with autoimmune endocrine disorders. The presence of IgG against the SARS-CoV-2 nucleocapsid in the absence of IgG against the SARS-CoV-2 spike RBD should be interpreted with caution.

## 1. Introduction

Severe acute respiratory syndrome coronavirus 2 (SARS-CoV-2) is a new infectious agent that causes coronavirus disease 2019 (COVID-19). Humoral immune responses play critical roles in protecting individuals against SARS-CoV-2 infection, particularly through the development of antibodies. There is an urgent need to understand humoral immune responses to SARS-CoV-2 and how these responses contribute to disease severity and vaccine-induced immunity. The reasons that COVID-19 ranges from asymptomatic to severe and fatal remain unknown. Studies have shown that the factors that can affect disease severity include preexisting immunity to seasonal human coronaviruses (HCoVs) [1], coinfection with other viruses [2], and the presence of autoantibodies against type I interferons (IFNs) [3].

There are currently five known human betacoronaviruses (β-CoVs; OC43, HKU1, severe acute respiratory syndrome coronavirus 1 (SARS-CoV-1), Middle East respiratory syndrome coronavirus (MERS-CoV), and SARS-CoV-2) and two human alphacoronaviruses (229E and NL63). The seasonal HCoVs 229E, OC43, NL63 and HKU1 may cause mild upper respiratory tract disease, and the pathogenic HCoVs SARS-CoV-1, MERS-CoV and SARS-CoV-2 are associated with severe respiratory disease. Although seasonal HCoVs are responsible for up to 15%–30% of common colds in adults, there are insufficient data on the epidemiology of these four viruses worldwide [4]. Cross reactions between preexisting antibodies against proteins of common seasonal coronaviruses and those of the new SARS-CoV-2 have been studied by several research groups. However, whether cross-reactive antibodies against seasonal coronaviruses protect against COVID-19 requires further study [5]. Some studies have shown that preexisting immunity to seasonal HCoVs and other respiratory viruses plays a limited role in the formation of SARS-CoV-2 humoral immune responses [6]. An association between the levels of antibodies against common cold HCoVs and the symptoms of COVID-19 has also been shown [7].

On the other hand, a confirmed COVID-19 diagnosis does not rule out coinfection with other viruses. The results of studies have shown that up to 20% of COVID-19 patients are coinfected with another respiratory virus [8]. The viral agents most frequently identified in patients hospitalized with pneumonia are rhinovirus, influenza virus, respiratory syncytial virus (RSV), parainfluenza virus (PIV), and adenovirus (AdV) [9]. IgM antibodies can be a sign of recent infection, as they are detected in the serum only for a couple of weeks in response to an infection, followed by class switching to IgG [10].

Type I IFNs constitute the main first line of defense against viruses. Bastard et al. recently found neutralizing autoantibodies against type I IFNs (IFN-α2 and IFN-ω) in 10% of patients with life-threatening COVID-19 pneumonia [3], and another study found these antibodies in 18% of patients with fatal COVID-19 [11].

The goal of this work was to design a unified assay to assess different factors possibly affecting the course of COVID-19. We developed a protein-based microarray to evaluate antiviral and anti-IFN antibodies in the serum of COVID-19 patients. The microarray contains antigens corresponding to betacoronaviruses (SARS-CoV-2, MERS-CoV, SARS-CoV-1, HKU1, and OC43), noncoronavirus respiratory viruses (influenza A (FluA), influenza B (FluB), PIV-1, PIV-2, PIV-3, AdV, and RSV), and type I IFNs (IFN-ω and IFN-α). Both IgG and IgM antibodies can be evaluated simultaneously in this test. Multiplex detection of antibodies in serum samples collected during the prepandemic and pandemic periods from different cohorts of patients and healthy donors allowed us to identify some IgG cross-reactivity between different coronaviruses and to confirm the importance of autoantibodies against type I IFNs as markers of severe COVID-19.

## 2. Materials and Methods

### 2.1 Clinical data and serum samples

#### Prepandemic cohort

The samples collected before May 2019 at the Endocrinology Research Centre, Ministry of Health of Russia, were included in the study as the negative control group. In total, 70 serum samples were collected from naive individuals (prepandemic negative control group): healthy persons (n=22) and patients with endocrine (autoimmune and nonautoimmune) pathology (n=48). The prepandemic cohort included patients with the following diseases: type 2 autoimmune polyglandular syndrome (APS) (n=13), type 1 APS (APS-1; n=6), autoimmune diabetes mellitus (n=7), Graves’ disease (n=5), rheumatoid arthritis (n=5), nonautoimmune diabetes mellitus (n=4), hypergonadotropic hypogonadism of autoimmune genesis (n=2), primary autoimmune adrenal insufficiency (n=2), nonautoimmune adrenal insufficiency (n=1), primary hyperparathyroidism (n=1), autoimmune thyroiditis (n=1), and adrenal lymphoma (n=1).

#### Pandemic cohort I—convalescent plasma donors

Convalescent plasma donor samples (n=103) were obtained from the Federal Research Clinical Center FMBA of Russia (Moscow). Anti-SARS-CoV-2 spike receptor-binding domain fragment Arg319-Phe541 (RBD) IgG was measured in these samples using both the developed microarray-based multiplex assay and ELISA (SARS-CoV-2-IgG-IFA test, Xema Ltd., Moscow, Russia).

#### Pandemic cohort II—vaccinated individuals and recovered COVID-19 patients

To assess the specificity of the anti-SARS-CoV-2 assay, serum samples from individuals vaccinated with the Sputnik V COVID-19 vaccine (Gamaleya National Research Centre for Epidemiology and Microbiology, Moscow, Russia) and patients who recovered from mild/moderate COVID-19 were included in the study (n=9). In this cohort, the time post vaccination with two doses/recovery from COVID-19 was 2-9 months.

#### Pandemic cohort III—patients with APS-1

To evaluate the detection of autoantibodies against type I IFNs using the developed assay, serum samples from APS-1 patients (n=9) were included in the study. Three patients had no history of COVID-19 or vaccination; six patients had recovered from mild/moderate COVID-19 and did not receive immunosuppressive therapy. For five of the APS-1 patients, paired samples were collected (prepandemic (2019) and during the pandemic (2021)).

#### Pandemic cohort IV— intensive care unit (ICU) patients

Samples collected from patients with severe COVID-19 confirmed with RT–PCR were included in the study as the positive controls. Serum samples were collected from 86 ICU patients. The samples were obtained from the Federal Research Clinical Center FMBA of Russia (Moscow).

### 2.2 Microarray manufacturing and multiplex assay for antibody detection

The following immobilized antigens were included the microarray layout (Figure 1): SARS-CoV-2 (spike RBD (S-RBD))—SARS-CoV-2 spike receptor-binding domain fragment Arg319-Phe541 (8COV1, HyTest, Finland), SARS-CoV-2 N—SARS-CoV-2 nucleocapsid (8COV3, HyTest, Finland), Flu-A—influenza A virus nucleoprotein (LS-G21636, LifeSpan BioSciences, USA), RSV—respiratory syncytial virus (8RSV79, HyTest, Finland), OC43 spike protein (DAGC131, Creative Diagnostics, USA), HKU1 spike protein (DAGC132, Creative Diagnostics, USA), IFN-ω—interferon ω (300-02J, PeproTech, USA); IFN-α-2a—interferon α-2a (11100-1, PBL Assay Science, USA), AdV—adenovirus (8AV13, HyTest, Finland), PIV-1—parainfluenza virus type 1 (8P76, HyTest, Finland), PIV-2—parainfluenza virus type 2 (8P76-2, HyTest, Finland), PIV-3—parainfluenza virus type 3 (8P76-3, HyTest, Finland), Flu-B—influenza B virus nucleoprotein (LS-G138331, LifeSpan BioSciences, USA), SARS-CoV (spike)—severe acute respiratory syndrome coronavirus spike protein (DAGC111, Creative Diagnostics, USA), and MERS-CoV (spike)—Middle East respiratory syndrome coronavirus spike protein (DAGH10300, Creative Diagnostics, USA).

**Figure 1.**
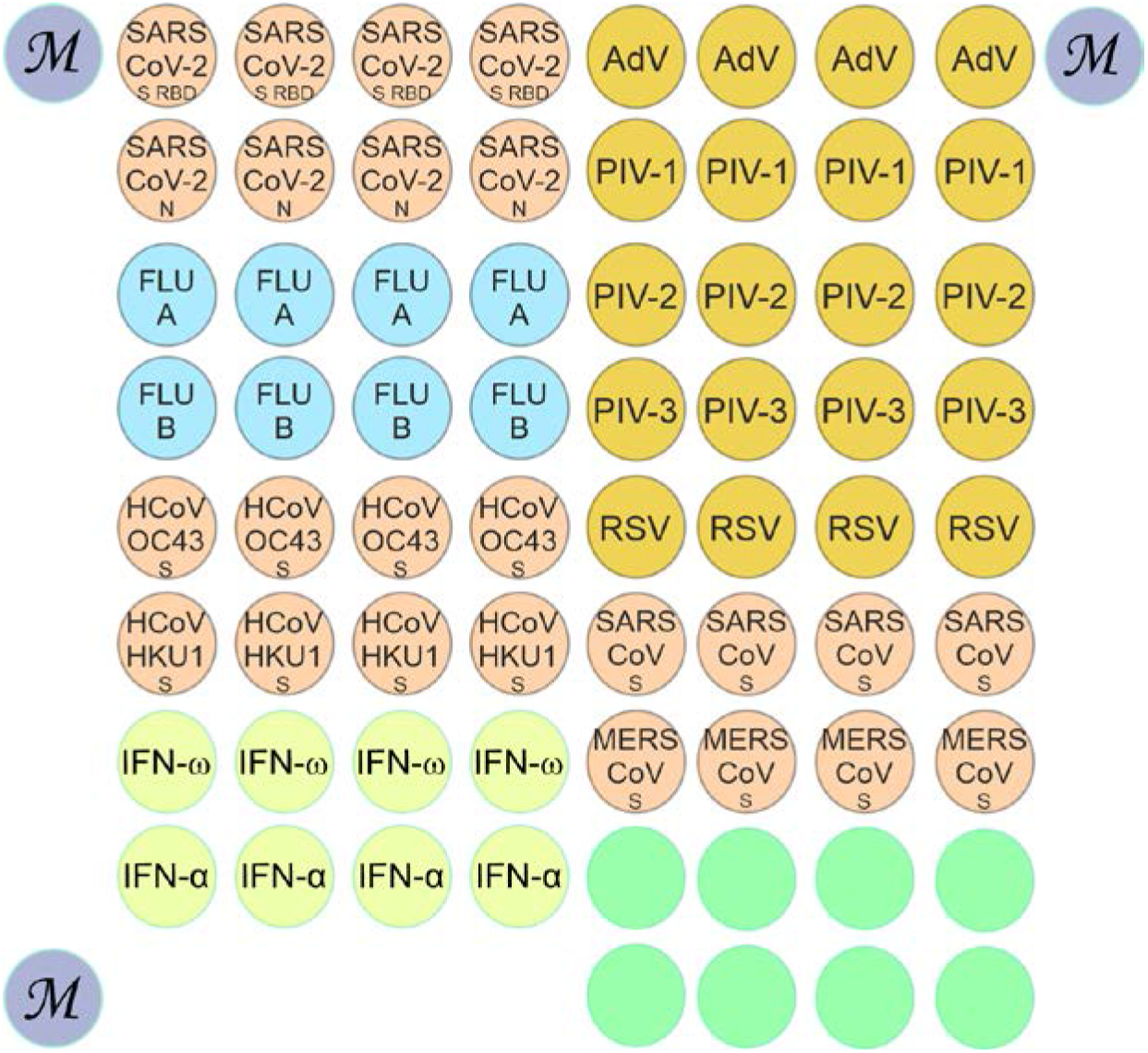
Configuration of the microarray for detecting antibodies against respiratory viruses and type I IFNs. Abbreviations: SARS-CoV-2 S RBD—SARS-CoV-2 spike receptor-binding domain fragment Arg319-Phe541, SARS-CoV-2 N—SARS-CoV-2 nucleocapsid, Flu A—influenza A virus nucleoprotein, Flu B—influenza B virus nucleoprotein, HCoV OC43 S—OC43 coronavirus spike protein, HCoV HKU1 S—HKU1 coronavirus spike protein, IFN-ω—interferon ω, IFN-α—interferon α-2a, AdV—adenovirus, PIV-1—parainfluenza virus type 1, PIV-2—parainfluenza virus type 2, PIV-3—parainfluenza virus type 3, RSV—respiratory syncytial virus, SARS-CoV S—severe acute respiratory syndrome coronavirus spike protein, MERS CoV S—Middle East respiratory syndrome coronavirus spike protein.

Molecular profiling of autoantibodies in serum samples was performed using hydrogel-based low-density microarrays [12]. Preparation of surfaces, mixture of gel monomers, and polymerization and blocking of microarrays were carried out as described earlier [13]. The diameter of the gel elements was 150 ± 20 µm, and the distance between the elements was 300 µm. Each antigen was immobilized in quadruplicate elements to improve the reproducibility of the assay results (Figure 1). The microarray consists of 71 hydrogel elements: 60 elements with immobilized viral proteins and IFNs, eight elements with the gel but no immobilized proteins and three elements with a fluorescent marker (M) to allow correct positioning of the microarray when acquiring a fluorescence image after analysis.

The microarrays were probed with human sera and analyzed using a modified version of a previously published technique [13]. In brief, blood serum samples from patients were diluted 1:100 in PBS buffer with 0.14% polyvinyl alcohol (50 kDa) and 0.14% polyvinylpyrrolidone (360 kDa) (all from Sigma–Aldrich, St. Louis, MO, USA) and applied to the microarray elements (100 µL). After incubation (usually overnight, 37°C), intermediate washing (PBS with 0.01% Tween 20 (Sigma–Aldrich, St. Louis, MO, USA), 20 min), rinsing and drying, the microarrays were treated with a mixture of fluorescently labeled antibodies (5 µg/mL; F(ab’) 2-goat anti-human IgG-Cy5, and 5 µg/mL, goat anti-IgM-Cy3; 50 µL in total) in PBS buffer with 0.14% polyvinyl alcohol, 0.14% polyvinylpyrrolidone and 1% BSA (Sigma–Aldrich, St. Louis, MO, USA). After incubation (30 min, 37°C), the microarrays were washed (PBS with 0.01% Tween 20, 30 min), rinsed with H2O, and dried. Anti-species antibodies (Invitrogen, Carlsbad, CA, USA) were preliminarily labeled with the N-hydroxysuccinimide esters Cy5 and Cy3 (GE Healthcare, Chicago, IL, USA) according to the manufacturer’s instructions, as indicated.

### 2.3 Analysis of fluorescence and interpretation of results

Fluorescence images of the microarrays were acquired using a laser-excited analyzer developed at the Engelhardt Institute of Molecular Biology (EIMB, Moscow, Russia) [14]. Measurement of microarray fluorescence was carried out using ImaGel Studio software (EIMB). For better reproducibility of each group (n) of four elements with the same antigen, the resulting In signal value was calculated as the median of the four corresponding fluorescence signal values [15]. The variation coefficient of the signals within one group of elements with the same antigen, i.e., for each data point, did not exceed 15%. In the group of eight elements without immobilized proteins, the resulting fluorescence signal value I_ref_ was calculated as the median of the eight corresponding fluorescence signal values. For anti-SARS-CoV-2 antibodies, I_n_/I_ref_ ratios for the corresponding groups of microarray elements were obtained upon analysis with samples from 22 prepandemic individuals, and three standard deviations above these ratios were taken as the cutoff values. For all other antibodies, I_n_/I_ref_ ratios for the corresponding groups were obtained upon analysis with buffer (PBS buffer with 0.14% polyvinyl alcohol and 0.14% polyvinylpyrrolidone; n= 10), and three standard deviations above these values were taken as the cutoff values. The cutoff values for selecting positive signals were I_n_/I_ref_ ≥2.2 for IgG antibodies and I_n_/I_ref_ ≥3.8 for IgM antibodies. Plots and correlation coefficients were generated with MedCalc Statistical Software version 20.008 (MedCalc Software bv, Ostend, Belgium; https://www.medcalc.org; 2020).

## 3. Results

### 3.1 Detection of antibodies with the microarray-based assay

To determine antibody titers in serum, a protein hydrogel microarray with immobilized target antigens was designed (Figure 1). The microarray contained antigens corresponding to respiratory viruses (SARS-CoV-2, SARS-CoV-1, MERS-CoV, OC43, HKU1; FluA and FluB; AdV; RSV; PIV-1, PIV-2, and PIV-3) and type I IFNs (IFN-ω and IFN-α-2a). Virus-specific IgG and IgM antibodies as well as autoantibodies against type I IFNs were assayed using diluted serum samples. To detect immunoglobulins bound to immobilized proteins, secondary anti-species antibodies labeled with two different fluorescent dyes (Cy5 and Cy3) were used. Figure 2 shows the fluorescence profiles of the microarray as the result of testing serum samples from patients with severe COVID-19.

**Figure 2.**
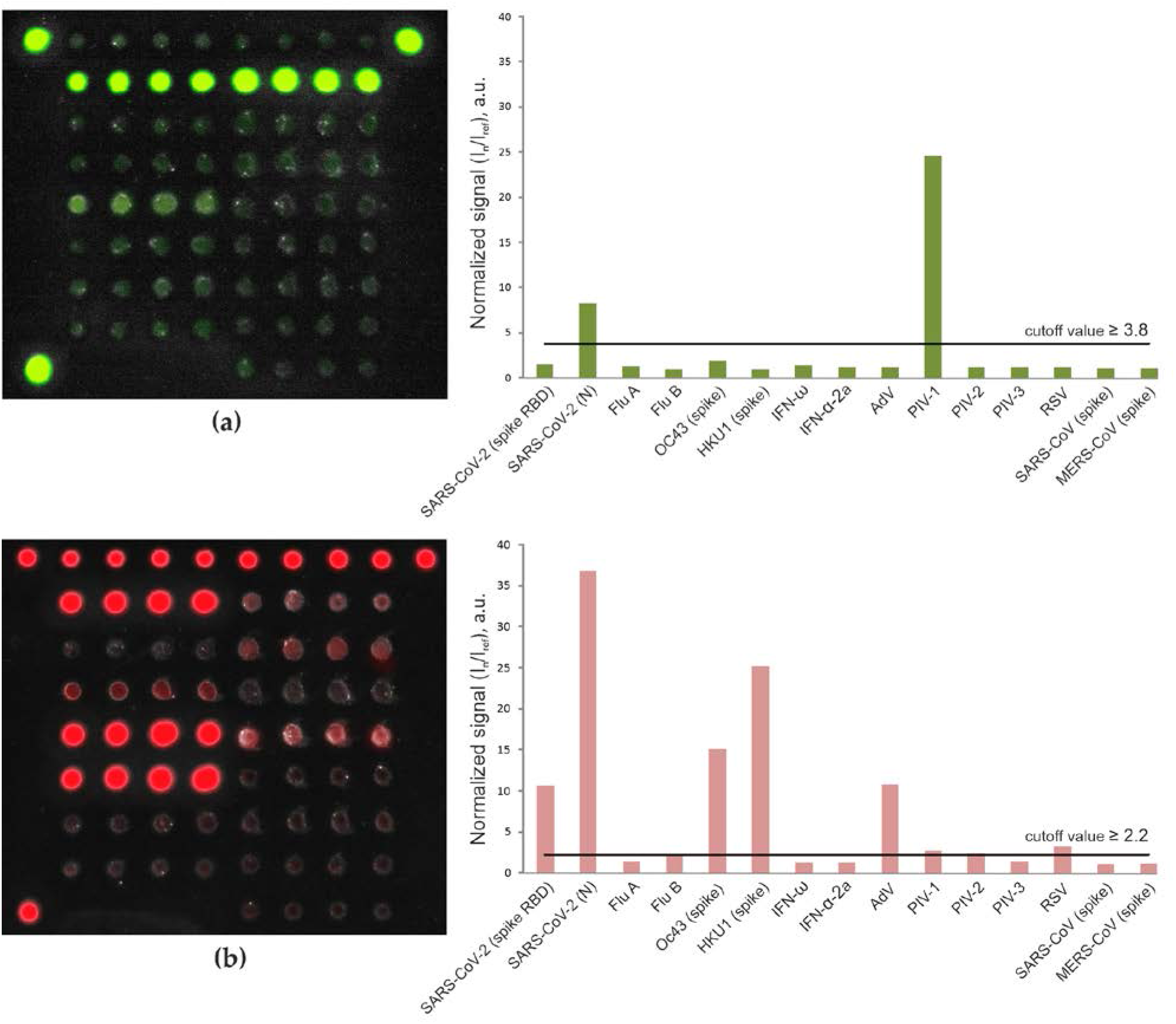
Fluorescence images of the microarray and values of normalized signals (I_n_/I_ref_) from the microarray elements after analysis of a serum sample from a COVID-19 ICU patient: IgM **(a)** and IgG **(b)** antibodies were detected with goat Cy3-conjugated anti-human IgM and Cy5-conjugated anti-human IgG. The cutoff values characterizing the positive signals of the microarray elements are shown by the horizontal line.

Analysis of the Cy3 fluorescence profile revealed two positive signals in the groups of elements containing SARS-CoV-2 nucleocapsid and PIV-1 (Figure 2a). Therefore, the tested sample was considered positive for IgM against these viruses. The Cy5 fluorescence profile was characterized by positive signals in the groups of elements containing SARS-CoV-2 spike RBD and nucleocapsid as well as those containing OC43 and HKU1 HCoV spike proteins, AdV, PIV-1 and RSV (Figure 2b). Thus, the microarray detected IgG antibodies against the corresponding viruses.

The concordance rate between the multiplex microarray-based assay and ELISA for the detection of IgG antibodies against the SARS-CoV-2 spike RBD was determined for 103 samples from convalescent plasma donors (pandemic cohort I). To calculate the accuracy, precision and recall, the positive and negative ELISA results were used as true positive and true negative results, respectively. The microarray results agreed well with the ELISA results, supporting the validity of the microarray as a serological assay in SARS-CoV-2 virus research (accuracy 85.4%, precision 1.0, recall 0.73). With the results for antibodies against any of the SARS-CoV-2 proteins detected by the microarray, the accuracy and recall increased to 95.1% and 0.91, respectively.

### 3.2 Detection of anti-SARS-CoV-2 antibodies in pandemic cohort II (patients with mild/moderate COVID-19 vs. vaccinated individuals)

To confirm assay specificity separately for anti-spike and anti-nucleocapsid antibodies, we included serum samples from patients who had recovered from mild/moderate COVID-19 (n = 5) and individuals who had been vaccinated with the Sputnik V COVID-19 vaccine (n = 4) in the study. All samples from the mild/moderate COVID-19 group were positive for IgG against both the SARS-CoV-2 spike and nucleocapsid, whereas samples from the vaccinated group were positive for IgG against only the SARS-CoV-2 spike RBD. None of the vaccinated patient sera demonstrated anti-SARS-CoV-2 nucleocapsid reactivity for either IgG or IgM antibodies (Figure 3).

**Figure 3.**
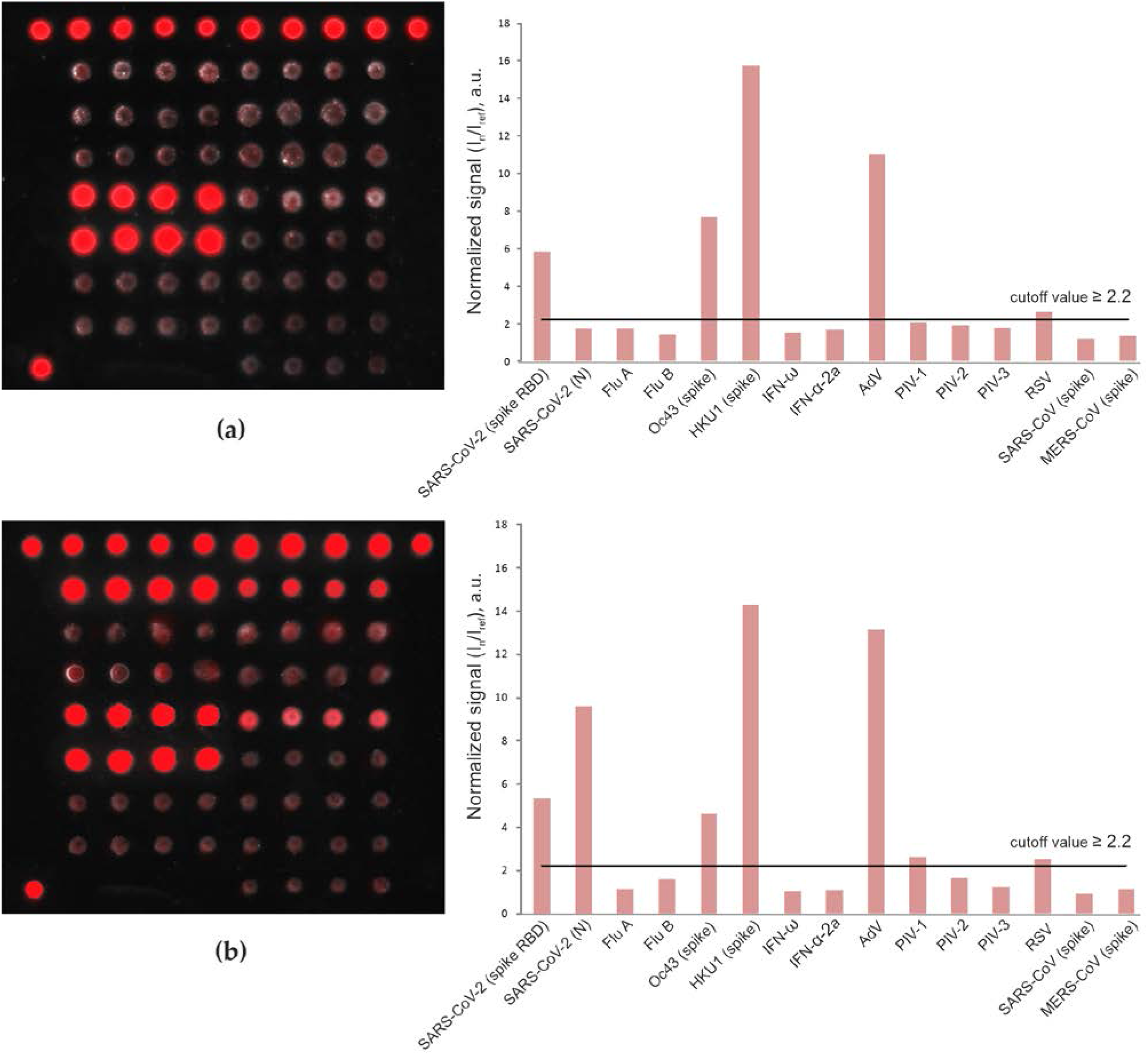
Fluorescence images of microarrays and values of normalized signals (I_n_/I_ref_) from the microarray elements after analysis of IgG in serum samples from a Sputnik V COVID-19 vaccine recipient **(a)** and a patient who had recovered from mild/moderate COVID-19 **(b)**.

### 3.3 Detection of IgG antibodies against betacoronaviruses

Most prepandemic and ICU patients had IgG antibodies against the seasonal HCoVs OC43 (spike) and HKU1 (spike) due to previous infections (Supplementary Table S1). In prepandemic serum, IgG against OC43 (spike) was detected in 68.6% (48/70) of the samples; in COVID-19 ICU patient serum, this percentage increased to 89.5% (77/86 samples). Similarly, in prepandemic serum, IgG against HKU1 (spike) was detected in 82.9% (58/70) of the samples; in COVID-19 ICU patient serum, this proportion increased to 88.4% (76/86 samples).

Figure 4 shows the distribution of the normalized fluorescence signals (I_n_/I_ref_) for IgG against betacoronaviruses obtained by microarray analysis in the group of COVID-19 ICU patients and in the control prepandemic cohort of healthy individuals and chronically ill patients.

**Figure 4.**
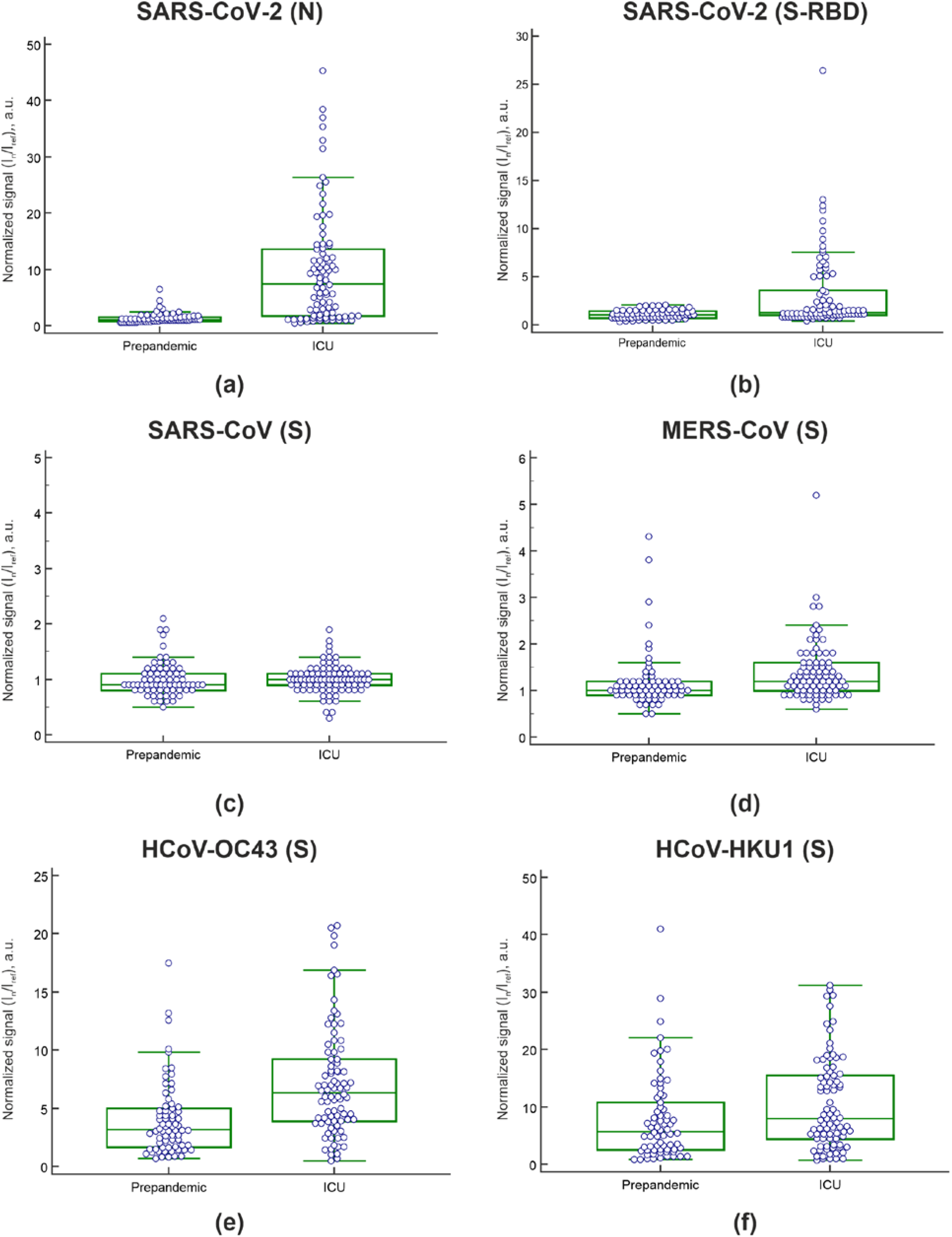
Humoral immune responses to betacoronaviruses in samples from the heterogeneous prepandemic cohort (n=70) and samples from COVID-19 ICU patients (n=86). **(a)** IgG against the SARS-CoV-2 nucleocapsid; **(b)** IgG against the SARS-CoV-2 spike RBD; **(c)** IgG against the SARS-CoV spike protein; **(d)** IgG against MERS-CoV spike protein; **(e)** IgG against the OC43 spike protein; **(f)** IgG against the HKU1 spike protein. The whisker plots show the distribution of IgG antibody titers across the prepandemic and ICU patient samples. The dots show the values of each sample. The solid black line indicates the median, and the top and bottom box boundaries indicate the first and third quartiles.

The cutoff value for antibodies against SARS-CoV-2 was determined using 22 prepandemic serum samples from healthy individuals collected before May 2019. For each antibody against SARS-CoV-2 proteins, samples with a fluorescence value greater than three standard deviations above the mean of the healthy prepandemic control samples were classified as positive. Six samples from the prepandemic cohort (n=48), which contained patients with various pathologies, including autoimmune endocrine disorders, were considered positive for IgG against the SARS-CoV-2 nucleocapsid, and no sample was considered positive for IgG against SARS-CoV-2 spike RBD. In these six samples, IgG against the seasonal coronaviruses OC43 (spike) and HKU1 (spike) were also detected. For three of these samples, positive signals were obtained for IgG MERS-CoV (spike). Two of the six samples were also positive for IgM against SARS-CoV-2 nucleocapsid. These antibodies were detected in the serum of patients with the following autoimmune diseases: autoimmune diabetes mellitus (n=2), Graves’ disease (n=2), and type 2 APS (n=2) (autoimmune diabetes mellitus + autoimmune thyroiditis, and autoimmune diabetes mellitus + autoimmune thyroiditis + Addison disease). Forty-three of the 48 prepandemic samples from patients with chronic diseases were previously characterized by ELISA and multiplex methods for positive organ-specific autoantibodies [13]. In each of the six samples positive for antibodies against the SARS-CoV-2, nucleocapsid organ-specific autoantibodies against one or more autoantigens were previously detected (Supplementary Table S2).

Correlations of the fluorescence levels for IgG antibodies against various betacoronaviruses were calculated using the values of fluorescence signals normalized to the reference signal from the empty gel elements (Table 1).

**Table 1.**
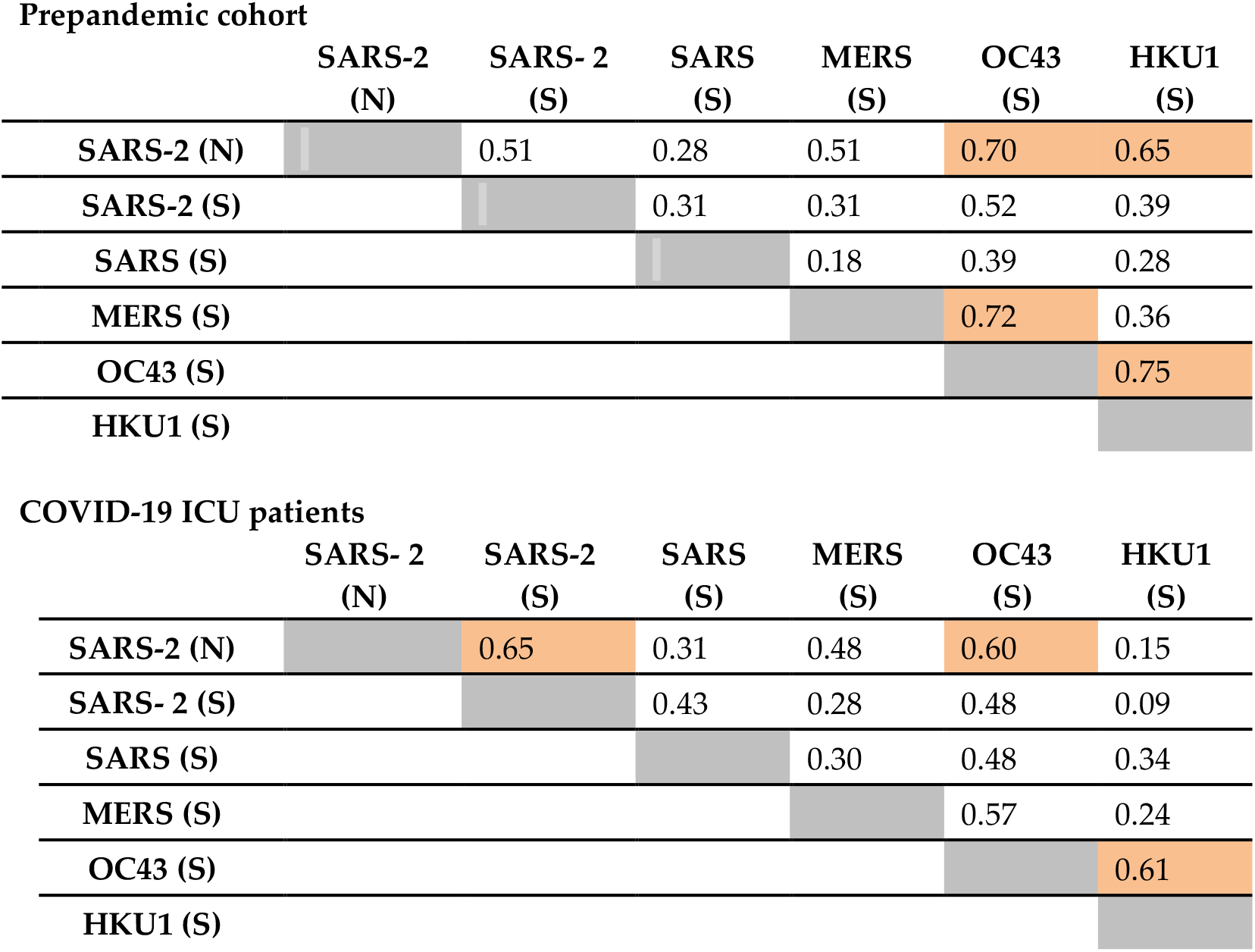
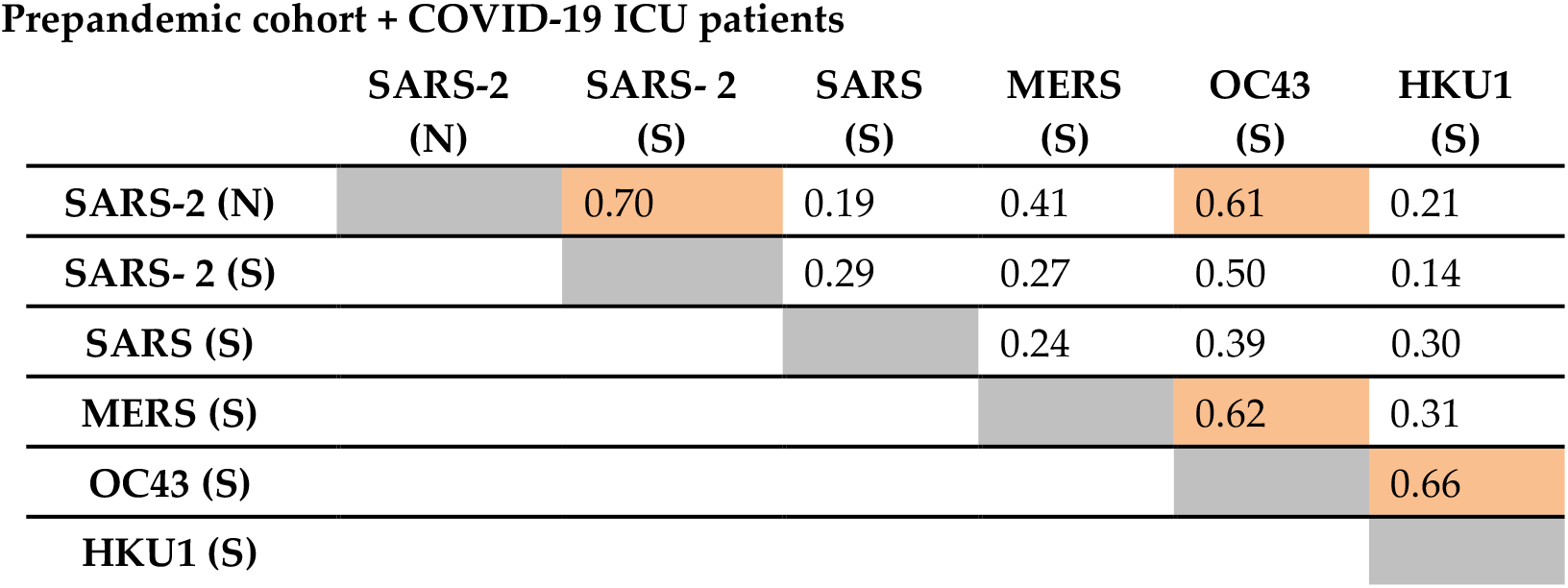
Correlation coefficients for IgG against different betacoronaviruses in the prepandemic (n=70), COVID-19 ICU (n=86), and integrated prepandemic + COVID-19 ICU (n=156) cohorts.

| Prepandemic cohort |  |  |  |  |  |  |
| --- | --- | --- | --- | --- | --- | --- |
|  | SARS-2<br>(N) | SARS- 2<br>(S) | SARS<br>(S) | MERS<br>(S) | OC43<br>(S) | HKU1<br>(S) |
| SARS-2 (N) |  | 0.51 | 0.28 | 0.51 | 0.70 | 0.65 |
| SARS-2 (S) |  |  | 0.31 | 0.31 | 0.52 | 0.39 |
| SARS (S) |  |  |  | 0.18 | 0.39 | 0.28 |
| MERS (S) |  |  |  |  | 0.72 | 0.36 |
| OC43 (S) |  |  |  |  |  | 0.75 |
| HKU1 (S) |  |  |  |  |  |  |
| COVID-19 ICU patients |  |  |  |  |  |  |
|  | SARS- 2<br>(N) | SARS-2<br>(S) | SARS<br>(S) | MERS<br>(S) | OC43<br>(S) | HKU1<br>(S) |
| SARS-2 (N) |  | 0.65 | 0.31 | 0.48 | 0.60 | 0.15 |
| SARS- 2 (S) |  |  | 0.43 | 0.28 | 0.48 | 0.09 |
| SARS (S) |  |  |  | 0.30 | 0.48 | 0.34 |
| MERS (S) |  |  |  |  | 0.57 | 0.24 |
| OC43 (S) |  |  |  |  |  | 0.61 |
| HKU1 (S) |  |  |  |  |  |  |

Prepandemic cohort + COVID-19 ICU patients
|  | SARS-2<br>(N) | SARS- 2<br>(S) | SARS<br>(S) | MERS<br>(S) | OC43<br>(S) | HKU1<br>(S) |
| --- | --- | --- | --- | --- | --- | --- |
| SARS-2 (N) |  | 0.70 | 0.19 | 0.41 | 0.61 | 0.21 |
| SARS- 2 (S) |  |  | 0.29 | 0.27 | 0.50 | 0.14 |
| SARS (S) |  |  |  | 0.24 | 0.39 | 0.30 |
| MERS (S) |  |  |  |  | 0.62 | 0.31 |
| OC43 (S) |  |  |  |  |  | 0.66 |
| HKU1 (S) |  |  |  |  |  |  |

### Prepandemic cohort + COVID-19 ICU patients

Figure 5 shows the correlations for IgG antibodies against the various betacoronaviruses included in the microarray in the prepandemic cohort of healthy individuals and chronically ill patients.

**Figure 5.**
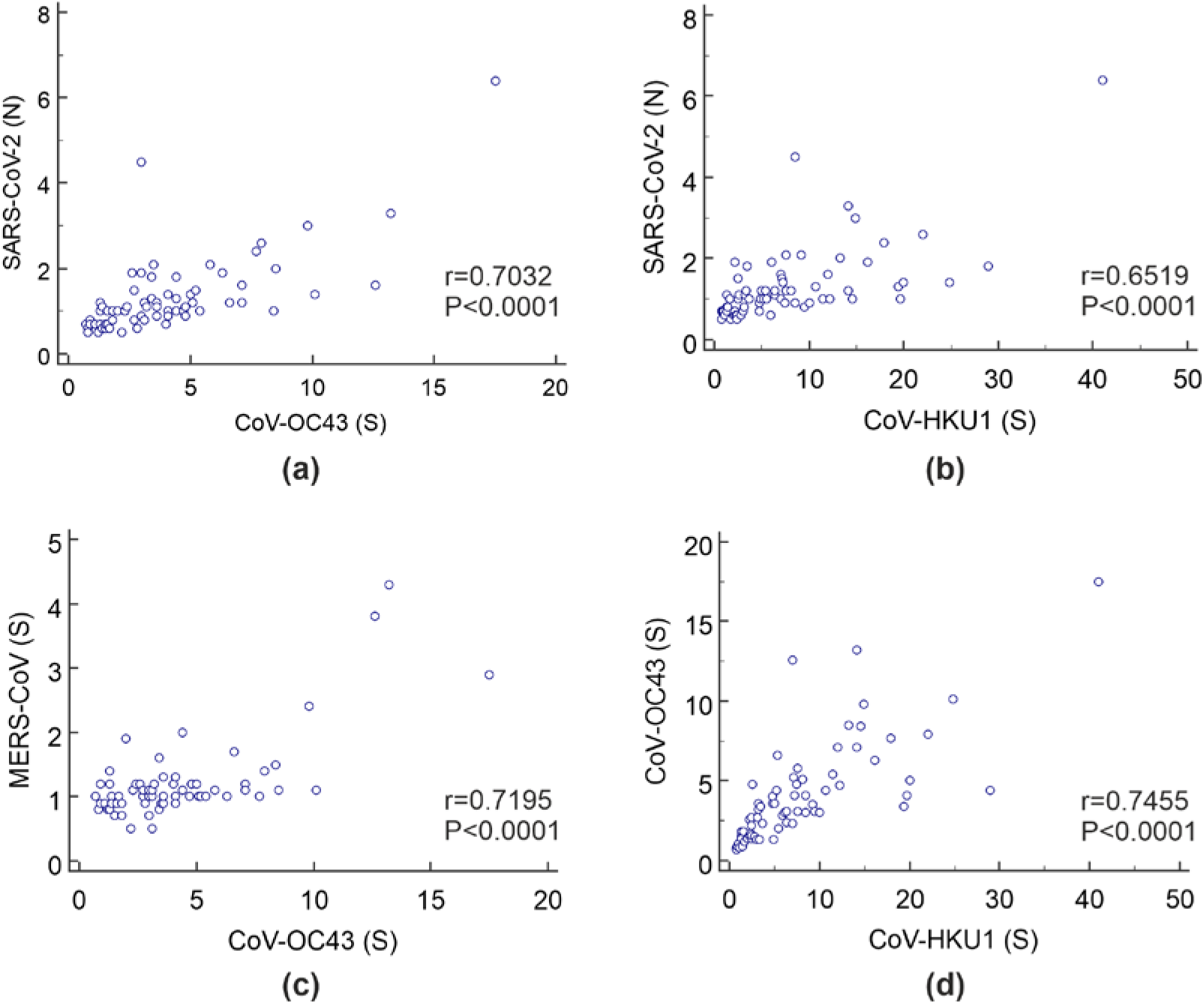
Scatter diagram for **(a)** IgG against SARS-CoV-2 (N) vs. IgG against OC43 (spike), **(b)** IgG against SARS-CoV-2 (N) vs. IgG against HKU1 (spike), **(c)** IgG against MERS-CoV (spike) vs. IgG against OC43 (spike), and **(d)** IgG against HKU1 (spike) vs. IgG against OC43 (spike) in the prepandemic cohort. The I_n_/I_ref_ values are plotted along the two axes.

In the naive prepandemic cohort, fairly strong positive correlations were observed between IgG against OC43 and HKU1 (r=0.75), IgG against OC43 and MERS-CoV (r=0.72), IgG against SARS-CoV-2 (N) and OC43 (r=0.70), and IgG against SARS-CoV-2 (N) and HKU1 (r=0.65). In addition, in the cohort of COVID-19 ICU patients, positive but comparatively weak correlations remained between IgG against OC43 and HKU1 (r=0.61) and IgG against SARS-CoV-2 (N) and OC43 (r=0.60).

### 3.4 Frequencies of antibodies against respiratory viruses

The proportions of positive IgG and IgM antibodies against all respiratory viruses included in the microarray in the prepandemic cohort and in COVID-19 ICU patients were determined using the developed assay (Figure 6). In the COVID-19 ICU patient cohort, the frequencies of antibodies against the seasonal coronaviruses OC43 and HKU1, AdV, RSV, PIV-1, and FluB were increased, and those against PIV-3 and FluA were decreased.

**Figure 6.**
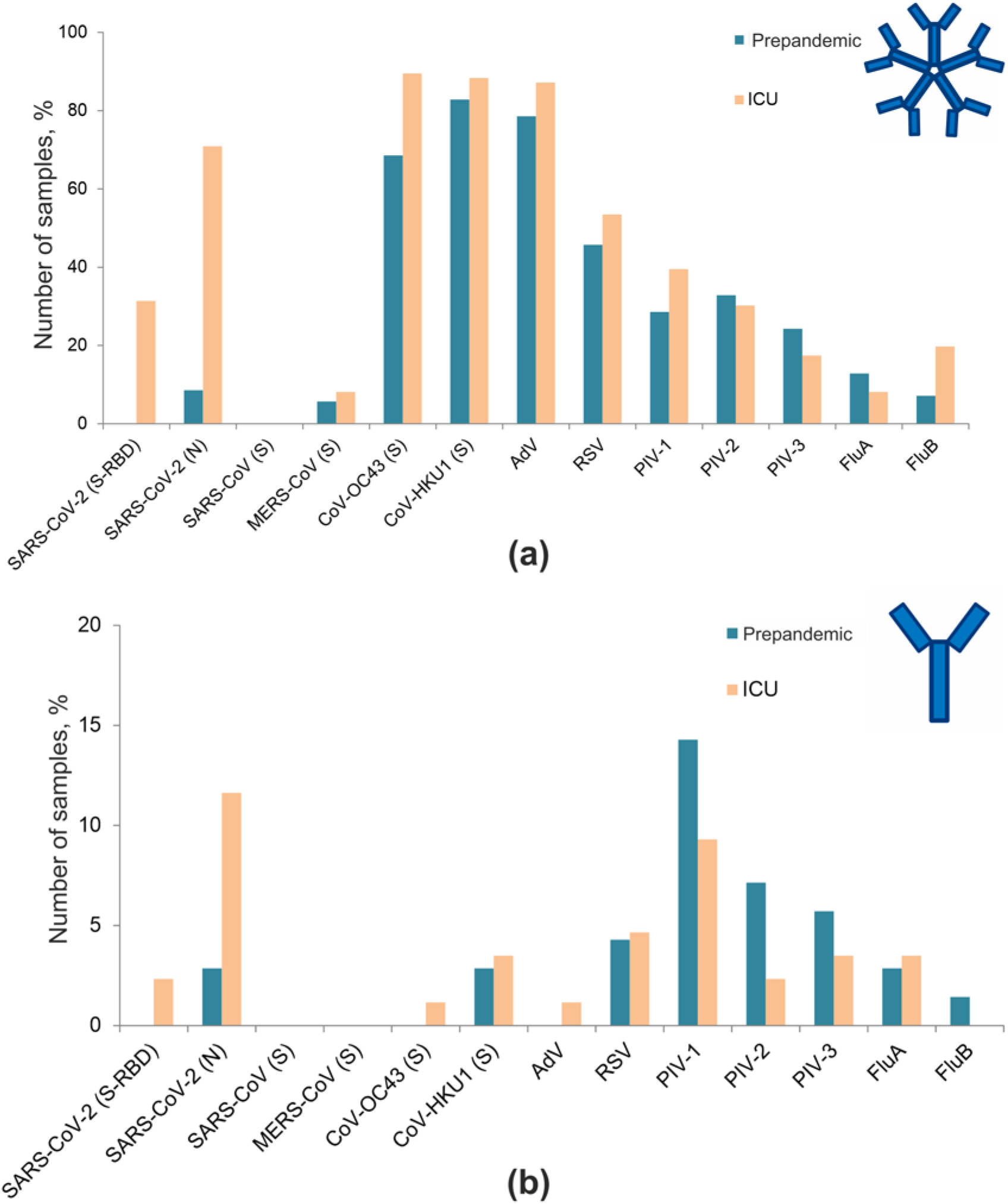
Frequencies of IgM **(a)** and IgG **(b)** antibodies against respiratory viruses in the prepandemic cohort and COVID-19 ICU patients.

### 3.5 Detection of autoantibodies against type I IFNs

APS-1 is characterized by high levels of autoantibodies (IgG) against IFN-ω and IFN-α [16], [13]. In addition, it has recently been shown that these antibodies can be markers for severe COVID-19 [3]. Figure 7 shows the distribution of IgG antibodies against type I IFNs in serum samples of APS-1 patients; in the prepandemic cohort, excluding patients with APS-1; and COVID-19 ICU patients. In 10.5% (9/86) of COVID-19 ICU patients, autoantibodies against type I IFNs were found. However, antibodies against both IFN-ω and IFN-α were detected in only three samples. Two serum samples contained only anti-IFN-ω antibodies, and four samples contained only anti-IFN-α antibodies. Moreover, only 4 of the 9 patients had anti-IFN antibody titers comparable to the high titers of these autoantibodies found in patients with APS-1. In one previously described patient with lymphoma in the prepandemic cohort, both anti-IFN-ω and anti-IFN-α antibodies were detected at high titers [13]. In two samples from healthy donors in the prepandemic cohort, anti-IFN antibodies were detected at rather low titers (I_n_/I_ref_ ratios of 2.5 and 3.3, respectively).

**Figure 7.**
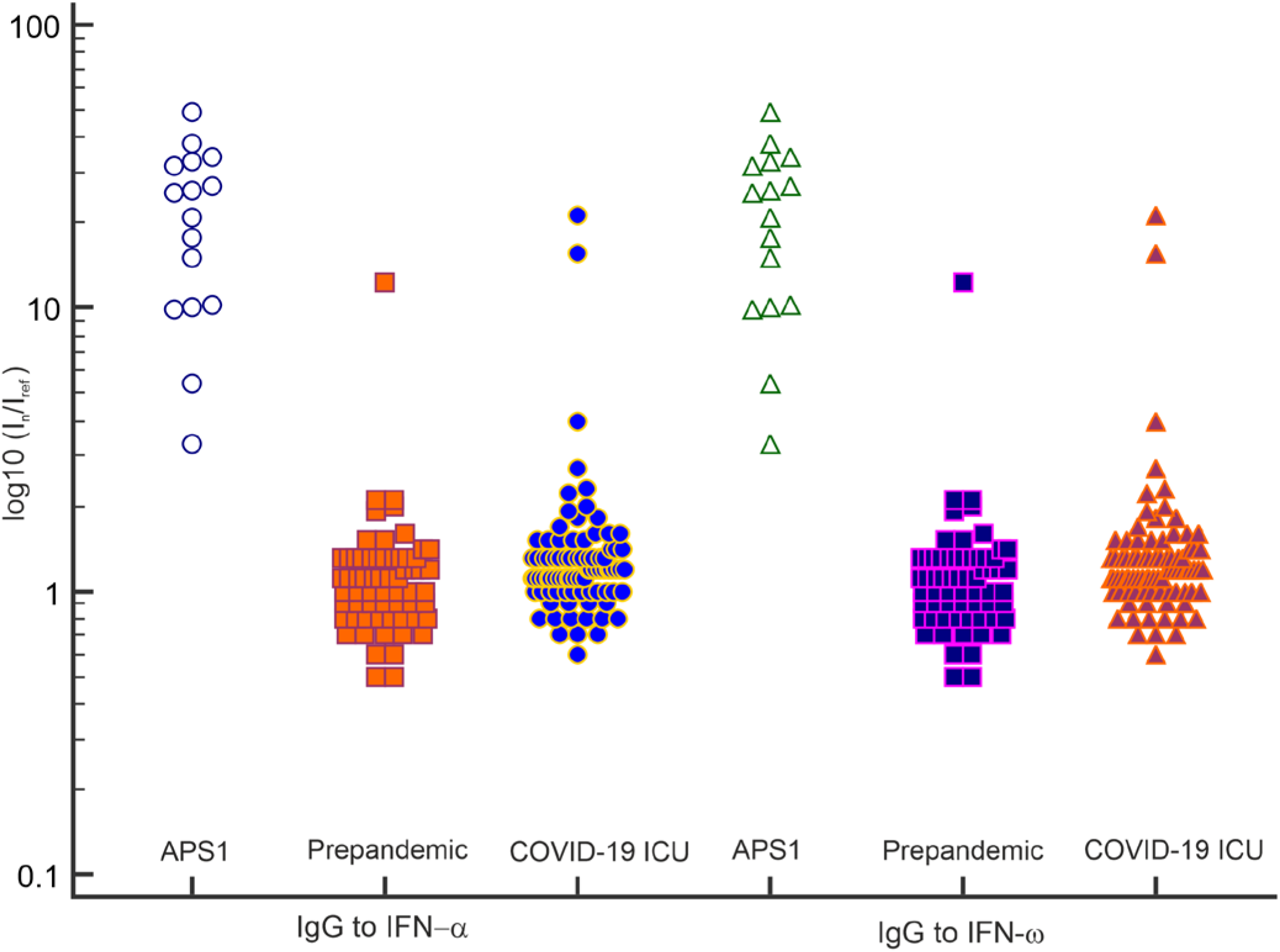
IgG antibodies against IFN-α and IFN-ω detected by the microarray assay in serum samples from APS-1 patients; the prepandemic cohort excluding APS-1 patients; and COVID-19 ICU patients. The values of the normalized fluorescence signals are log10-transformed.

A total of 17 samples from 12 APS-1 patients were included in the study. Seven samples were collected before the pandemic in 2019; ten were collected in 2021. Paired serum samples collected in 2019 and 2021 were available for five patients (Supplementary Figure S1). There were similarly high levels of IgG against IFN-α and IFN-ω in both the prepandemic and 2021 APS-1 patient samples. However, in the paired sera (n=5), the signal level was decreased for APS-1 patients who were infected with SARS-CoV-2 (n=4); this decrease was not observed in the paired sera from the one patient not exposed to SARS-CoV-2. In the assay comparing the serum samples of APS-1 patients after recovery from COVID-19 with the prepandemic samples from these patients, noticeable decreases in the signal for IgG against IFN-ω were observed: 32.8 (2019) vs. 27.0 (2021), 25.3 (2019) vs. 5.4 (2021), 48.8 (2019) vs. 31.5 (2021), and 31.1 (2019) vs. 7.1 (2021). For IgG against IFN-α, the corresponding decreases were 50.5 (2019) vs. 37.4 (2021), 53.6 (2019) vs. 31.9 (2021), 61.1 (2019) vs. 31.7 (2021), and 45.3 (2019) vs. 27.2 (2021).

None of the prepandemic sera from APS-1 patients (n=7) or pandemic sera from patients not exposed to SARS-CoV-2 (n=3) showed reactivity above the cutoff value for either IgG or IgM against SARS-CoV-2 antigens.

## 4. Discussion

Multiplex analysis of SARS-CoV-2-specific antibodies is a very attractive idea; therefore, numerous microarrays have been developed to date [17–20]. In this study, we describe the development of a multiplex assay based on a low-density hydrogel microarray [12]. The assay detects antibodies against SARS-CoV-2 and other respiratory viruses as well as antibodies against type I IFNs. The test can simultaneously detect IgG and IgM antibodies against human betacoronaviruses (OC43, HKU1, SARS-CoV-1, MERS-CoV, and SARS-CoV-2); RSV; AdV; PIV-1, PIV-2, and PIV-3; FluA and FluB; and type I IFNs (α and ω).

The developed multiplex assay was applied to track the cross-reactivity of antibodies targeting SARS-CoV-2, to detect changes in the IgG repertoire after SARS-CoV-2 infection or vaccination, to estimate the incidence of IgM antibodies against other respiratory viruses, and to estimate the percentage of patients in a cohort of patients with severe COVID-19 with autoantibodies against type I IFNs.

Differences in vaccine-induced immunity and natural infection-induced humoral immunity allowed us to demonstrate the specificity of the assay for the SARS-CoV-2 spike and nucleocapsid. Sputnik V vaccine recipients generated antibody responses to the spike protein, while patients who recovered after natural infection generated a wide range of antibodies against viral proteins, including the spike and nucleocapsid. As shown in this small sample, the microarray assay can unambiguously distinguish vaccine recipients from recovered COVID-19 patients (Figure 3).

Generally, the microarray results and the ELISA results exhibited good concordance. However, detection of IgG against the SARS-CoV-2 spike RBD was less accurate than simultaneous detection of antibodies against both the spike RBD and nucleocapsid. The decreased frequencies of spike RBD-reactive antibodies compared to nucleocapsid antibodies in the COVID-19 ICU patient cohort can be explained by several factors. First, the time elapsed since the onset of COVID-19 was unknown for the studied ICU cohort. Since IgM and IgG antibody levels change over time after infection, this parameter can be critical. Second, we used the antigenic RBD region of the SARS-CoV-2 spike protein (Arg319-Phe541 fragment). Antibody responses against the RBD have been shown to be weaker than those against the S1, S2 and N proteins; moreover, detection of the RBD by antibodies does not parallel the detection of S1 [21]. Thus, further increases in the sensitivity and specificity of the microarray can be achieved by immobilizing not only the spike RBD and nucleocapsid but also a number of other SARS-CoV-2 proteins, including other spike subunits, membrane proteins, etc.

Most prepandemic samples as well as samples from COVID-19 ICU patients contained IgG against the spike protein of the OC43 and HKU1 coronaviruses. Studies have revealed that, the number of cross-reactive antibodies that recognize seasonal HCoV antigens is increased in patients infected with SARS-CoV-2 [22]. We showed an increase in IgG against OC43 and a less pronounced increase in IgG against HKU1 in the cohort of COVID-19 ICU patients compared with the naive prepandemic cohort.

To assess the diagnostic accuracy of the microarray in a population that had not been exposed to SARS-CoV-2, serum samples collected before May 2019 were used. Since the results of antibody assays for patients with chronic inflammatory diseases can be misleading, a heterogeneous group of patients, a naive prepandemic control group was used as a negative control. False-positive results for IgG antibodies against the SARS-CoV-2 nucleocapsid were detected in prepandemic serum samples of patients with autoimmune endocrine diseases. Six of the 48 prepandemic samples from chronically ill patients produced false-positive results for IgG against the SARS-CoV-2 nucleocapsid. Among these patients were patients with type 1 diabetes, autoimmune thyroid diseases, and type 2 APS. None of the prepandemic samples from healthy individuals or chronically ill patients without autoimmune disorders contained antibodies against the SARS-CoV-2 nucleocapsid. Four of the prepandemic samples from chronically ill patients and seven samples from COVID-19 ICU patients also showed binding of IgG against the MERS-CoV spike, to which it is unlikely that these individuals had been exposed (Figure 4d).

Positive SARS-CoV-2 antibody test results in naive populations have previously been identified in stored prepandemic sera [10], patients with chronic inflammatory diseases [23], and uninfected individuals [24]. The possible causes of false-positive detection of SARS-CoV-2 antibodies can be endogenous interfering factors, including rheumatoid factor, heterophile antibodies, human anti-animal antibodies, lysozyme, complement, and cross antigens, or exogenous interfering factors, including incomplete coagulation, specimen contamination, and insufficient optimization of the diagnostic kit’s reaction system [25]. For antibodies against viruses, cross-species interactions can have an influence. Cross-reactivity of immune responses between SARS-CoV-2 and seasonal coronavirus has recently been reported [26–28]. As shown by Tso et al., the SARS-CoV-2 nucleocapsid is the major antigen recognized by these cross-reactive antibodies [29]. In our study, in the naive prepandemic cohort, cross-reactivity was observed between antibodies against the SARS-CoV-2 nucleocapsid and the spike proteins of the OC43 and HKU1 seasonal coronaviruses, between antibodies against the OC43 spike and the HKU1 spike, and between antibodies against the OC43 spike and the MERS-CoV spike. However, it remains unclear whether the false-positive signals that we observed were the result of cross-reactive interactions with antibodies against other coronaviruses and whether the analysis result was influenced by endogenous factors associated with an autoimmune inflammatory process or by a combination of factors. Antibodies against the nucleocapsid of SARS-CoV-2 are an attractive diagnostic marker, as they appear earlier than antibodies against the spike protein [30]. However, considering possible cross reactions, the ideal choice for the development of an immunoassay is simultaneous assessment of both the nucleocapsid and spike proteins.

Consistent with a recent study by Xu et al, similar epidemiological data for the frequencies of IgG antibodies against AdV and RSV were obtained [31]. However, the expected high prevalence of IgG antibodies against the FluA and FluB nucleoproteins was not observed. Most samples were found to contain IgG against OC43 (spike) and IgG against HKU1 (spike), consistent with previously published data [32]. Moreover, IgG against OC43 (spike) and IgG against HKU1 (spike) were codetected in 78.2% of patients. IgG antibodies against three types of human PIVs (PIV-1, PIV-2 and PIV-3) were codetected in 10.9% of patients. Half of the samples (50.6%) were found to contain IgG antibodies against at least one PIV. Thus, a high prevalence of IgG antibodies against respiratory viruses, which usually cause mild illness with upper respiratory symptoms in nonimmunodeficient adults, was found in the studied cohort.

As the early symptoms of COVID-19 are similar to those of other acute respiratory viral infections, a multiplex assay to detect IgM antibodies against a wide range of respiratory viruses may be helpful to reveal possible coinfections. Although IgM antibodies can be a sign of recent infection, IgM assays have limitations. First, IgM antibodies are released only at specific time points. In addition, they have lower affinity for viral antigens than IgG antibodies. Thus, the performance of IgM assays can vary and demonstrate low sensitivity, as shown in different studies [33]. In the studied cohort of COVID-19 ICU patients, IgM-class antibodies against more than two different viruses were found simultaneously in 10.5% of the samples.

Anti-type I IFN antibodies are highly specific for APS-1 patients [16]. In addition, pre-existing autoantibodies against IFN-α and IFN-ω can be prognostic markers for severe COVID-19 [3]. The specificity of a microarray assay for anti-type I IFN antibodies was previously confirmed for a slightly different assay [13]. In this study, the presence of autoantibodies against type I IFNs in the cohort of critically ill COVID-19 ICU patients was detected in 10.5% of the samples.

Using serum samples from APS-1 patients with known recent exposure to SARS-CoV-2, we tried to assess the changes in the levels of autoantibodies against type I IFNs in these patients compared with unexposed patients. All included APS-1 patients who contracted SARS-CoV-2 developed only mild / moderate COVID-19 symptoms and did not receive immunosuppressive therapy. Consistent with previous studies [34], similarly high levels of antibodies against type I IFNs were detected in the prepandemic (2019) and 2021 samples. However, four of the paired samples from APS-1 patients showed a notable decrease in the anti-type I IFN antibody level in the post COVID-19 recovery sample compared to the paired prepandemic sample. In contrast, the paired serum samples from the only patient with APS-1 not exposed to SARS-CoV-2 did not show the same trend. The developed method is not intended to quantify the levels of autoantibodies, and the sample size is too small to draw a conclusion. However, given the very low prevalence of APS-1 patients in the population, we can assume that this observed trend cannot be overlooked, and further research is needed. As type I IFNs are at the first line of the innate immune response to viral infection, it can be speculated that during COVID-19, some preexisting anti-type I IFN antibodies interact with their targets as the level of IFNs increases. The formation of these circulating immune complexes can lead to decreased levels of autoantibodies.

## 5. Conclusions

Advanced serologic testing is essential to understand the spread of and immunity to SARS-CoV-2. The presence of cross-reactive antibodies in samples can potentially affect assay specificity, causing false-positive results. Given the high prevalence of antibodies against seasonal coronaviruses in the population, these cross reactions need to be evaluated during the validation of SARS-CoV-2 assays. Our findings indicate that the results of assays designed to detect antibodies against the SARS-CoV-2 nucleocapsid protein should be interpreted with caution, especially in patients with autoimmune diseases. A result of anti-SARS-CoV-2 nucleocapsid IgG-positive and anti-SARS-CoV-2 spike IgG-negative should be interpreted as truly positive for current or previous COVID-19 only at specific time points after symptom onset.

## Data Availability

All data produced in the present work are contained in the manuscript and supplementary

## Supplementary Materials

The following are available online at www.mdpi.com/xxx/s1, Figure S1: APS-1 patient’s samples collected in 2019 and 2021, Table S1: I_n_/I_ref_ ratios obtained by microarray for all samples, Table S2: Prepandemic samples of patients with chronic diseases.

## Author Contributions

E.S. designed the project, performed the experiments and wrote the manuscript; M.F. realized microarray manufacturing and performed the experiments; V.V-E. obtained ELISA data; N.N., M.Yu and E.T. prepared serum samples for prepandemic cohort and pandemic cohort III; V.B. prepared serum samples for pandemic cohorts I and IV; A.I. prepared serum samples for pandemic cohort II, D.G. supervised the project and wrote the manuscript. All authors have read and agreed to the published version of the manuscript.

## Institutional Review Board Statement

According to the Ethics Committees of the Endocrinology Research Centre, Ministry of Health of Russia, Moscow, Russia, Federal Research Clinical Center FMBA of Russia, Moscow, this research does not require ethical approval. All specimens used in this study were anonymous samples that omitted personal information about the patients, particularly their name or address.

## Informed Consent Statement

Informed consent was obtained from all subjects involved in the study.

## Data Availability Statement

The authors confirm that the data supporting the findings of this study are available within the article and/or its supplementary materials.

## References

1. Kaplonek, P.; Wang, C.; Bartsch, Y.; Fischinger, S.; Gorman, M.J.; Bowman, K.; Kang, J.; Dayal, D.; Martin, P.; Nowak, R.P.; et al. Early cross-coronavirus reactive signatures of humoral immunity against COVID-19. Science Immunology 2021, 6, 1–13, doi:10.1126/sciimmunol.abj2901.

2. Burrel, S.; Hausfater, P.; Dres, M.; Pourcher, V.; Luyt, C.E.; Teyssou, E.; Soulié, C.; Calvez, V.; Marcelin, A.G.; Boutolleau, D. Co-infection of SARS-CoV-2 with other respiratory viruses and performance of lower respiratory tract samples for the diagnosis of COVID-19. International Journal of Infectious Diseases 2021, 102, 10–13, doi:10.1016/j.ijid.2020.10.040.

3. Bastard, P.; Rosen, L.B.; Zhang, Q.; Michailidis, E.; Hoffmann, H.H.; Zhang, Y.; Dorgham, K.; Philippot, Q.; Rosain, J.; Béziat, V.; et al. Autoantibodies against type I IFNs in patients with life-threatening COVID-19. Science (New York, N.Y.) 2020, 370, doi:10.1126/science.abd4585.

4. Liu, D.X.; Liang, J.Q.; Fung, T.S. Human Coronavirus-229E, -OC43, -NL63, and -HKU1 (Coronaviridae). In Encyclopedia of Virology; Elsevier, 2021; pp. 428–440 ISBN 9780128096338.

5. Zedan, H.T.; Nasrallah, G.K. Is preexisting immunity to seasonal coronaviruses limited to cross-reactivity with SARS-CoV-2? A seroprevalence cross-sectional study in north-eastern France. EBioMedicine 2021, 71, 103580, doi:10.1016/j.ebiom.2021.103580.

6. Loos, C.; Atyeo, C.; Fischinger, S.; Burke, J.; Slein, M.D.; Streeck, H.; Lauffenburger, D.; Ryan, E.T.; Charles, R.C.; Alter, G. Evolution of Early SARS-CoV-2 and Cross-Coronavirus Immunity. mSphere 2020, 5, 1–10, doi:10.1128/msphere.00622-20.

7. Ortega, N.; Ribes, M.; Vidal, M.; Rubio, R.; Aguilar, R.; Williams, S.; Barrios, D.; Alonso, S.; Hernández-Luis, P.; Mitchell, R.A.; et al. Seven-month kinetics of SARS-CoV-2 antibodies and role of pre-existing antibodies to human coronaviruses. Nature Communications 2021, 12, 1–10, doi:10.1038/s41467-021-24979-9.

8. Kim, D.; Quinn, J.; Pinsky, B.; Shah, N.H.; Brown, I. Rates of Co-infection between SARS-CoV-2 and Other Respiratory Pathogens. JAMA - Journal of the American Medical Association 2020, 323, 2085–2086, doi:10.1001/jama.2020.6266.

9. Ruuskanen, O.; Lahti, E.; Jennings, L.C.; Murdoch, D.R. Viral pneumonia. The Lancet 2011, 377, 1264–1275, doi:10.1016/S0140-6736(10)61459-6.

10. Latiano, A.; Tavano, F.; Panza, A.; Palmieri, O.; Niro, G.A.; Andriulli, N.; Latiano, T.; Corritore, G.; Gioffreda, D.; Gentile, A.; et al. False-positive results of SARS-CoV-2 IgM/IgG antibody tests in sera stored before the 2020 pandemic in Italy. International Journal of Infectious Diseases 2021, 104, 159–163, doi:10.1016/j.ijid.2020.12.067.

11. Bastard, P.; Gervais, A.; Le Voyer, T.; Rosain, J.; Philippot, Q.; Manry, J.; Michailidis, E.; Hoffmann, H.-H.; Eto, S.; Garcia-Prat, M.; et al. Autoantibodies neutralizing type I IFNs are present in ∼4% of uninfected individuals over 70 years old and account for ∼20% of COVID-19 deaths. Science Immunology 2021, 6, eabl4340, doi:10.1126/sciimmunol.abl4340.

12. Gryadunov, D.A.; Shaskolskiy, B.L.; Nasedkina, T. V.; Rubina, A.Y.; Zasedatelev, A.S. The EIMB Hydrogel Microarray Technology: Thirty Years Later. Acta Naturae 2018, 10, 4–18, doi:10.32607/20758251-2018-10-4-4-18.

13. Savvateeva, E.N.; Yukina, M.Y.; Nuralieva, N.F.; Filippova, M.A.; Gryadunov, D.A.; Troshina, E.A. Multiplex Autoantibody Detection in Patients with Autoimmune Polyglandular Syndromes. International Journal of Molecular Sciences 2021, 22, 5502, doi:10.3390/ijms22115502.

14. Lysov, Y.; Barsky, V.; Urasov, D.; Urasov, R.; Cherepanov, A.; Mamaev, D.; Yegorov, Y.; Chudinov, A.; Surzhikov, S.; Rubina, A.; et al. Microarray analyzer based on wide field fluorescent microscopy with laser illumination and a device for speckle suppression. Biomedical Optics Express 2017, 8, 4798, doi:10.1364/boe.8.004798.

15. Rubina, A.Y.; Filippova, M.A.; Feizkhanova, G.U.; Shepeliakovskaya, A.O.; Sidina, E.I.; Boziev, K.M.; Laman, A.G.; Brovko, F.A.; Vertiev, Y. V.; Zasedatelev, A.S.; et al. Simultaneous detection of seven staphylococcal enterotoxins: Development of hydrogel biochips for analytical and practical application. Analytical Chemistry 2010, 82, 8881–8889, doi:10.1021/ac1016634.

16. Meager, A.; Visvalingam, K.; Peterson, P.; Möll, K.; Murumägi, A.; Krohn, K.; Eskelin, P.; Perheentupa, J.; Husebye, E.; Kadota, Y.; et al. Anti-interferon autoantibodies in autoimmune polyendocrinopathy syndrome type 1. PLoS Medicine 2006, 3, 1152–1164, doi:10.1371/journal.pmed.0030289.

17. Gillot, C.; Douxfils, J.; Cadrobbi, J.; Laffineur, K.; Dogné, J.-M.; Elsen, M.; Eucher, C.; Melchionda, S.; Modaffarri, É.; Tré-Hardy, M.; et al. An Original ELISA-Based Multiplex Method for the Simultaneous Detection of 5 SARS-CoV-2 IgG Antibodies Directed against Different Antigens. Journal of Clinical Medicine 2020, 9, 3752, doi:10.3390/jcm9113752.

18. Brynjolfsson, S.F.; Sigurgrimsdottir, H.; Einarsdottir, E.D.; Bjornsdottir, G.A.; Armannsdottir, B.; Baldvinsdottir, G.E.; Bjarnason, A.; Gudlaugsson, O.; Gudmundsson, S.; Sigurdardottir, S.T.; et al. Detailed Multiplex Analysis of SARS-CoV-2 Specific Antibodies in COVID-19 Disease. Frontiers in Immunology 2021, 12, 1–6, doi:10.3389/fimmu.2021.695230.

19. de Assis, R.R.; Jain, A.; Nakajima, R.; Jasinskas, A.; Felgner, J.; Obiero, J.M.; Norris, P.J.; Stone, M.; Simmons, G.; Bagri, A.; et al. Analysis of SARS-CoV-2 antibodies in COVID-19 convalescent blood using a coronavirus antigen microarray. Nature Communications 2021, 12, doi:10.1038/s41467-020-20095-2.

20. van Tol, S.; Mögling, R.; Li, W.; Godeke, G.J.; Swart, A.; Bergmans, B.; Brandenburg, A.; Kremer, K.; Murk, J.L.; van Beek, J.; et al. Accurate serology for SARS-CoV-2 and common human coronaviruses using a multiplex approach. Emerging Microbes and Infections 2020, 9, 1965–1973, doi:10.1080/22221751.2020.1813636.

21. Shah, J.; Liu, S.; Potula, H.; Bhargava, P.; Cruz, I.; Force, D.; Bazerbashi, A.; Ramasamy, R. IgG and IgM antibody formation to spike and nucleocapsid proteins in COVID-19 characterized by multiplex immunoblot assays. BMC Infectious Diseases 2021, 21, 325, doi:10.1186/s12879-021-06031-9.

22. Woudenberg, T.; Pelleau, S.; Anna, F.; Attia, M.; Donnadieu, F.; Gravet, A.; Lohmann, C.; Seraphin, H.; Guiheneuf, R.; Delamare, C.; et al. Humoral immunity to SARS-CoV-2 and seasonal coronaviruses in children and adults in north-eastern France. EBioMedicine 2021, 70, doi:10.1016/j.ebiom.2021.103495.

23. Kharlamova, N.; Dunn, N.; Bedri, S.K.; Jerling, S.; Almgren, M.; Faustini, F.; Gunnarsson, I.; Rönnelid, J.; Pullerits, R.; Gjertsson, I.; et al. False Positive Results in SARS-CoV-2 Serological Tests for Samples From Patients With Chronic Inflammatory Diseases. Frontiers in Immunology 2021, 12, 1–11, doi:10.3389/fimmu.2021.666114.

24. Ng, K.W.; Faulkner, N.; Cornish, G.H.; Rosa, A.; Harvey, R.; Hussain, S.; Ulferts, R.; Earl, C.; Wrobel, A.G.; Benton, D.J.; et al. Preexisting and de novo humoral immunity to SARS-CoV-2 in humans. Science 2020, 370, 1339–1343, doi:10.1126/science.abe1107.

25. Ye, Q.; Zhang, T.; Lu, D. Potential false-positive reasons for SARS-CoV-2 antibody testing and its solution. Journal of Medical Virology 2021, 93, 4242–4246, doi:10.1002/jmv.26937.

26. Sealy, R.E.; Hurwitz, J.L. Cross-reactive immune responses toward the common cold human coronaviruses and severe acute respiratory syndrome coronavirus 2 (Sars-cov-2): Mini-review and a murine study. Microorganisms 2021, 9, doi:10.3390/microorganisms9081643.

27. Hicks, J.; Klumpp-Thomas, C.; Kalish, H.; Shunmugavel, A.; Mehalko, J.; Denson, J.P.; Snead, K.R.; Drew, M.; Corbett, K.S.; Graham, B.S.; et al. Serologic Cross-Reactivity of SARS-CoV-2 with Endemic and Seasonal Betacoronaviruses. Journal of Clinical Immunology 2021, 41, 906–913, doi:10.1007/s10875-021-00997-6.

28. Ladner, J.T.; Henson, S.N.; Boyle, A.S.; Engelbrektson, A.L.; Fink, Z.W.; Rahee, F.; D’ambrozio, J.; Schaecher, K.E.; Stone, M.; Dong, W.; et al. Epitope-resolved profiling of the SARS-CoV-2 antibody response identifies cross-reactivity with endemic human coronaviruses. Cell Reports Medicine 2021, 2, doi:10.1016/j.xcrm.2020.100189.

29. Tso, F.Y.; Lidenge, S.J.; Peña, P.B.; Clegg, A.A.; Ngowi, J.R.; Mwaiselage, J.; Ngalamika, O.; Julius, P.; West, J.T.; Wood, C. High prevalence of pre-existing serological cross-reactivity against severe acute respiratory syndrome coronavirus-2 (SARS-CoV-2) in sub-Saharan Africa. International Journal of Infectious Diseases 2021, 102, 577–583, doi:10.1016/j.ijid.2020.10.104.

30. Keaney, D.; Whelan, S.; Finn, K.; Lucey, B. Misdiagnosis of SARS-CoV-2: A Critical Review of the Influence of Sampling and Clinical Detection Methods. Medical Sciences 2021, 9, 36, doi:10.3390/medsci9020036.

31. Xu, G.J.; Kula, T.; Xu, Q.; Li, M.Z.; Vernon, S.D.; Ndung’u, T.; Ruxrungtham, K.; Sanchez, J.; Brander, C.; Chung, R.T.; et al. Comprehensive serological profiling of human populations using a synthetic human virome. Science 2015, 348, aaa0698–aaa0698, doi:10.1126/science.aaa0698.

32. Gorse, G.J.; Patel, G.B.; Vitale, J.N.; O’Connor, T.Z. Prevalence of antibodies to four human coronaviruses is lower in nasal secretions than in serum. Clinical and Vaccine Immunology 2010, 17, 1875–1880, doi:10.1128/CVI.00278-10.

33. Al-Jighefee, H.T.; Yassine, H.M.; Al-Nesf, M.A.; Hssain, A.A.; Taleb, S.; Mohamed, A.S.; Maatoug, H.; Mohamedali, M.; Nasrallah, G.K. Evaluation of Antibody Response in Symptomatic and Asymptomatic COVID-19 Patients and Diagnostic Assessment of New IgM/IgG ELISA Kits. Pathogens 2021, 10, 161, doi:10.3390/pathogens10020161.

34. Bastard, P.; Orlova, E.; Sozaeva, L.; Lévy, R.; James, A.; Schmitt, M.M.; Ochoa, S.; Kareva, M.; Rodina, Y.; Gervais, A.; et al. Preexisting autoantibodies to type I IFNs underlie critical COVID-19 pneumonia in patients with APS-1. The Journal of experimental medicine 2021, 218, doi:10.1084/jem.20210554.

