## Supplementary figures and images for "Microarray-based detection of antibodies against SARS-CoV-2 proteins, common respiratory viruses and type I interferons"

### Figure S1

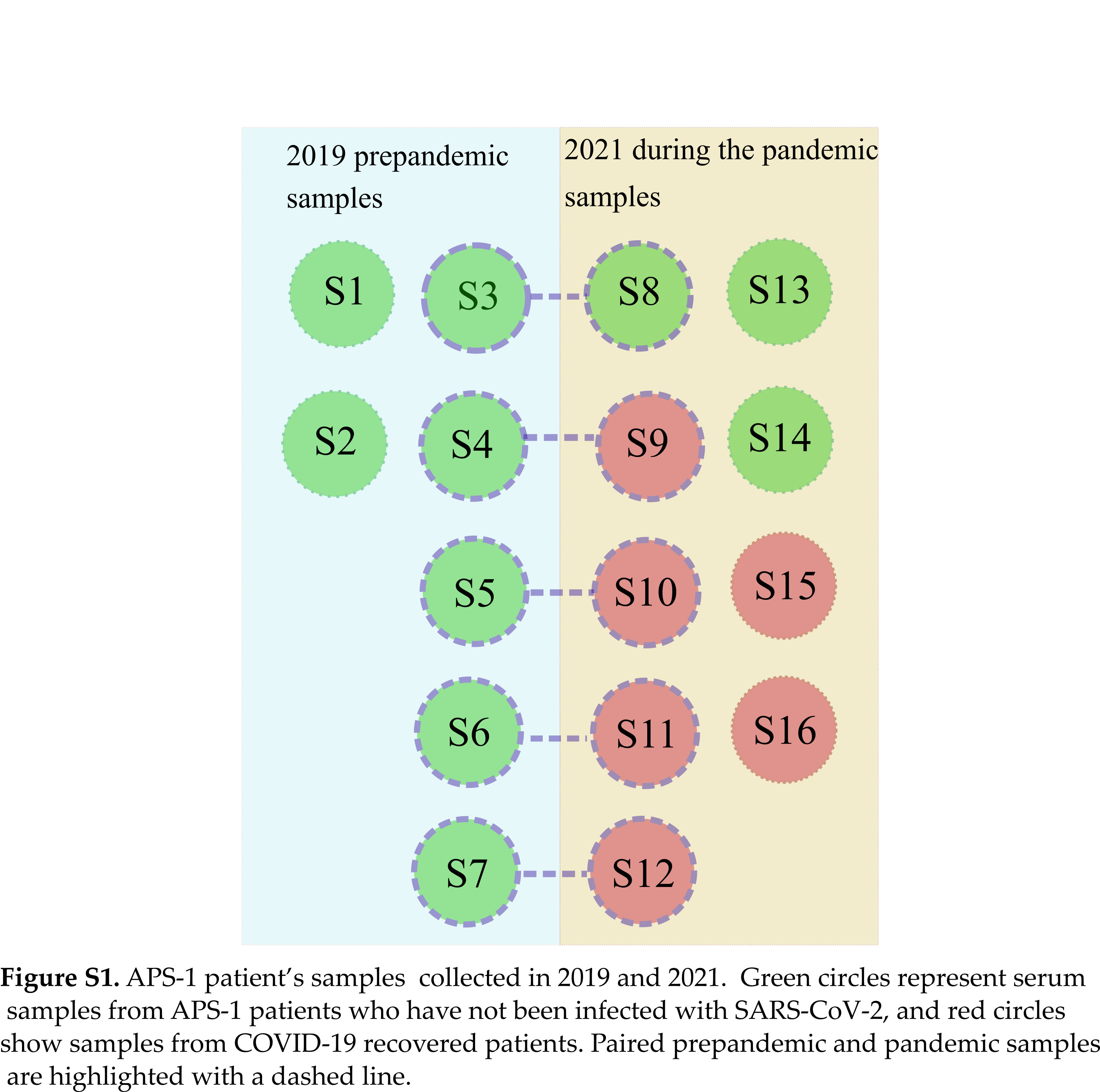
